# Laboratory-based surveillance of COVID-19 in the Greater Helsinki area, Finland, February-June 2020

**DOI:** 10.1101/2020.07.03.20145615

**Authors:** H Jarva, M Lappalainen, O Luomala, P Jokela, AE Jääskeläinen, AJ Jääskeläinen, H Kallio-Kokko, E Kekäläinen, L Mannonen, H Soini, S Suuronen, A Toivonen, C Savolainen-Kopra, R Loginov, S Kurkela

## Abstract

Laboratory registry data (80,791 specimens, 70,517 individuals) was used to characterise age- and sex-specific SARS-CoV-2 RT-PCR sampling frequency and positivity rate, and laboratory capacity building in Greater Helsinki, Finland during February-June 2020. While the number of positive cases was similar in males and females, the positivity rate was significantly higher in males. The highest incidence/100,000 was observed in those aged ≥80 years. The proportion of young adults in positive cases increased in late May 2020.

---

The WHO has advocated for the “test, trace, treat” strategy in the mitigation of COVID-19 pandemic [1]. Rarely has sophisticated laboratory diagnostics been set up at this pace and extent. As the epidemiological situation evolved quickly, the laboratory capacity building over the past few months played a key role in the epidemic response in each country. At the same time, laboratory-based surveillance can provide high quality data for public health management. By using laboratory registry data, the aim of this study was to characterise age- and sex-specific sampling frequency and positivity rate, and to characterise laboratory capacity building of SARS-CoV-2 RT-PCR testing in Greater Helsinki area in Finland during February-June 2020.

## Registry data

This study was based on the laboratory registry database of the Helsinki University Hospital Laboratory (HUS Diagnostic Center, HUSLAB), Finland. This laboratory provides services to Greater Helsinki, with a population of 1,685,983 (48.8% males, 51.2% females) as per 31 December 2019 [2].

The data included the date of sampling, sex, age, and the SARS-CoV-2 RT-PCR test result on specimens from 1 February to 15 June 2020. The data were collected according to permit HUS/157/2020 (Helsinki University Hospital, Finland). The data were analysed with GraphPad Prism^®^ according to tests and individual cases. In the case analysis, only the first test of the negative cases was counted. For the positive cases, only the first positive test was counted.

Data from the National Infectious Disease Register of the National Institute for Health and Welfare (THL) were retrieved to calculate age-specific incidence.

## Laboratory methods

The respiratory specimens were subjected to one of the following methods: a protocol based on Corman et al. [3], cobas® SARS-CoV-2 test kit on the cobas® 6800 system (Roche Diagnostics, Basel, Switzerland), Amplidiag® COVID-19 test (Mobidiag, Espoo, Finland) and Mobidiag Novodiag® Covid-19 assay (Mobidiag, Espoo, Finland). The performance of these tests in our laboratory is reported elsewhere [4, 5].

## Results

Of the 86,927 specimens sent to HUSLAB for SARS-CoV-2 RT-PCR testing between 1 February and 15 June 5,061 were excluded as they originated outside of Greater Helsinki. Additional 1,075 specimens were excluded as they gave repeatedly an invalid test result, or were never analysed due to a preanalytical failure. The final data included 80,791 specimens (Figure 1a) from 70,517 individuals: 29,885 (37.0%) specimens from 25,948 individual males, and 50,906 (63.0%) specimens from 44,569 individual females. The tested individuals had a median age of 43 years, mean 45 years, and a range of 0 days - 106 years. Altogether 4,057/80,791 (5.0%) of the specimens were positive, and 3,915/70,517 (5.6%) of the individuals were found positive.

**Figure 1.**
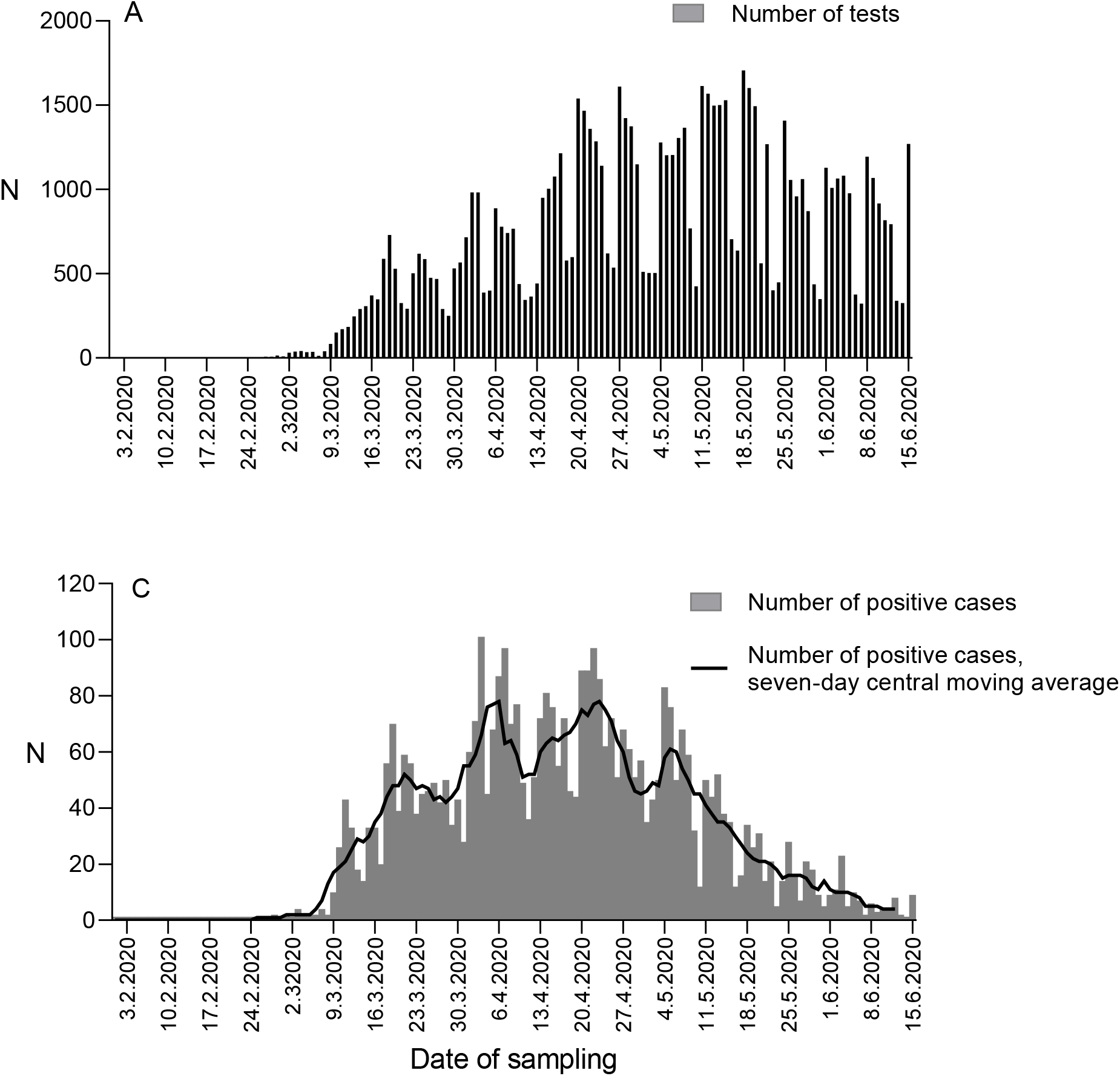
A) The number of SARS-CoV-2 RT-PCR tests conducted in HUSLAB by date of sampling, between 1 March and 15 June, 2020. B) The number of SARS-CoV-2 positive cases (columns) with seven-day central moving average (line) by date of sampling, between 1 March and 15 June, 2020.

The first positive case in Greater Helsinki was diagnosed on 25 February (Figure 1b). This was the second COVID-19 case diagnosed in Finland, the first being a Chinese tourist in Lapland [6]. As of 9 March, both the number of tests, positive tests and cases increased rapidly (Figure 1). A peak was reached on 3 April (96 new cases) (Figure 1b), and the daily number of new cases remained high throughout April and early May.

During the epidemic peak, the daily number of tests was still increasing through rapid capacity building. The daily number of tests (by sampling date) increased from 308 tests on March 15 to 1005 tests on April 15, and further to 1530 tests on May 15 (Figure 1a).

The highest proportion of positive cases was found in the age group 50-59 years (6.7%), and the lowest in the age group 0-9 years (2.9%) (Figure 2a). The number of test positive cases was highest in working-age adults (20-59 years) (Figure 2b). The age stratification of new positive cases over time showed a shift towards an increasing proportion of young adults (20-39 years) as of late May (Figure 2c). The incidence per 100,000 population in the Greater Helsinki area was 287/100,000 population, and highest in those aged ≥80 years (560/100,000) (Figure 2d).

**Figure 2.**
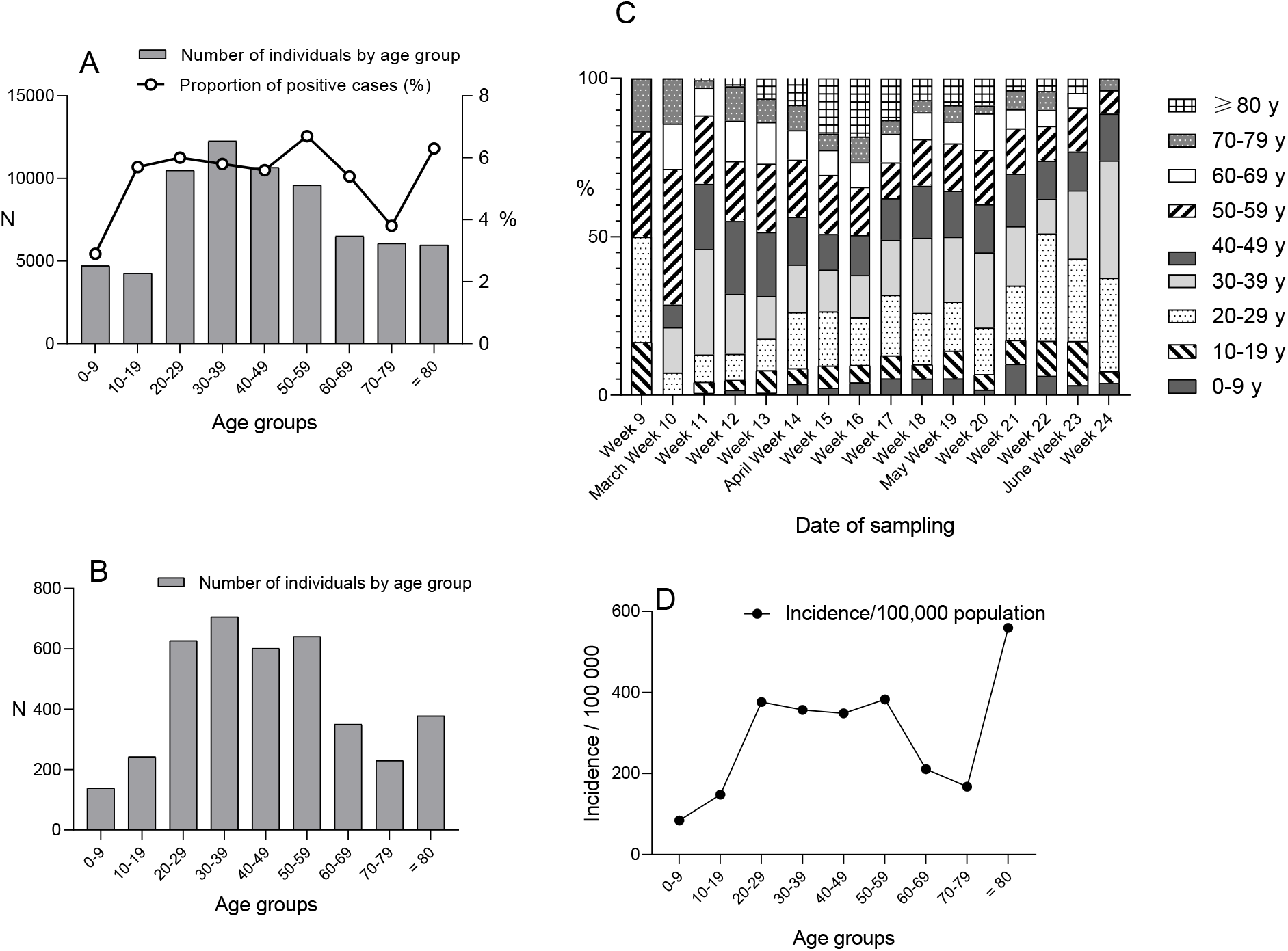
A) The number of individuals tested for SARS-CoV-2 RT-PCR by age group (columns) and the proportion of SARS-CoV-2 positive cases (line). B) The number of SARS-CoV-2 positive cases by age group. C) Age stratification of SARS-CoV-2 positive cases by calendar week of 2020. The proportion (%) of each age group is shown. D) Age-specific incidence/100,000 population of SARS-CoV-2 in the Greater Helsinki area; data from the National Infectious Diseases Registry.

In age group 0-29 years, the proportion of test positive cases peaked in weeks 13-15, while in age group 30-79 years the highest proportions were observed already in weeks 11 and 12 (Figure 3). In those aged ≥80 years, the proportion of positive cases began to increase in week 13, and peaked in week 15 (Figure 3).

**Figure 3.**
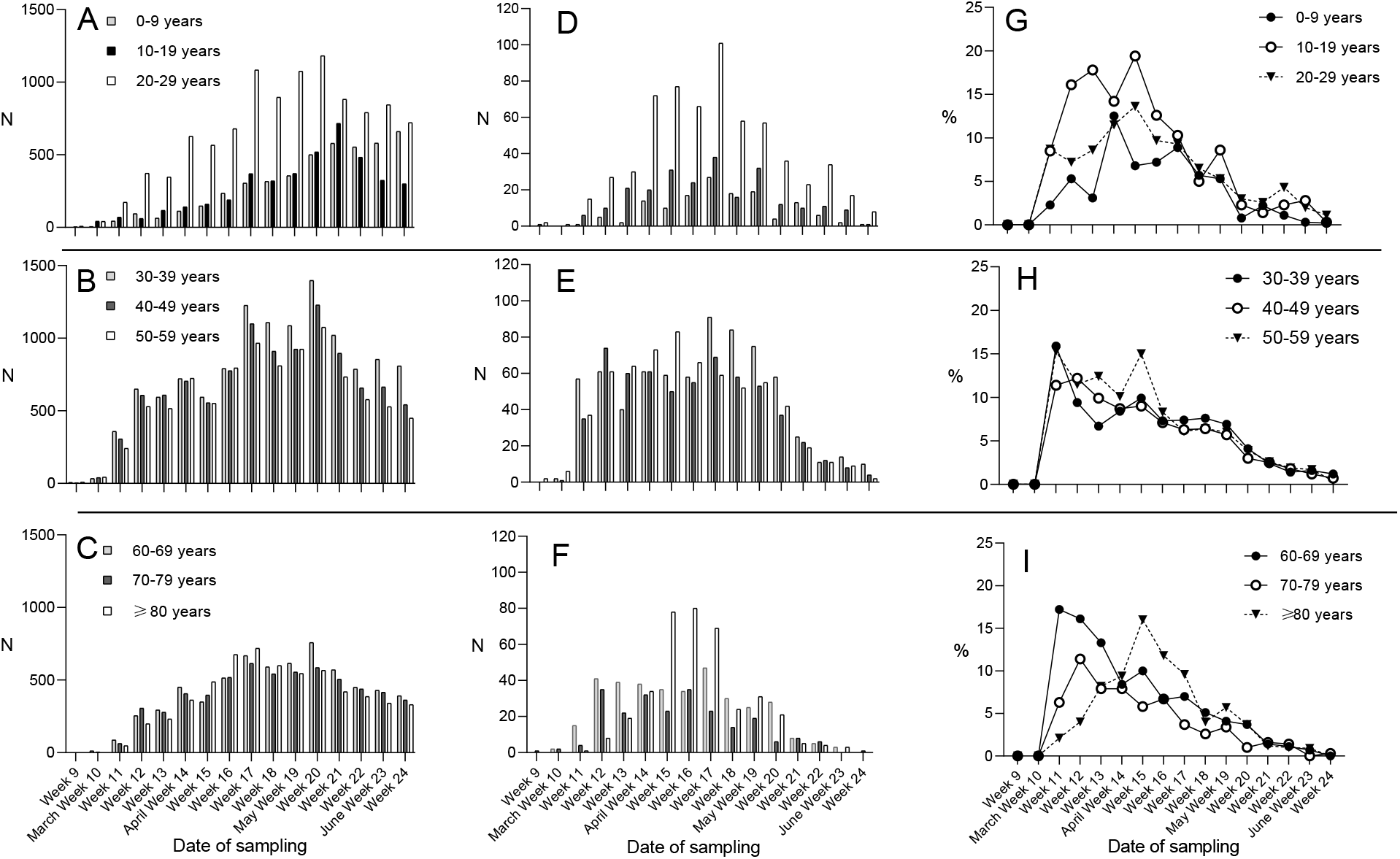
A-C) The age-specific number of individuals tested for SARS-CoV-2 RT-PCR by calendar week of 2020. D-F) The age-specific number of SARS-CoV-2 positive cases by calendar week of 2020. G-I) The age-specific proportion (%) of SARS-CoV-2 positive cases by calendar week of 2020.The age range for all tested males was 1 d – 102 years (mean 43.1 and median 41 years) and for positive males 2 months – 97 years (mean 43.5 and median 41 years). The age range for all tested females was 0 d – 106 years (mean 45.5 and median 43 years) and for positive females 1 month – 100 years (mean 47.7 and median 45 years).

Of the positive cases, 1935/3,915 (49.4%) were males, and 1980/3,915 (50.6%) females, and they had a median age of 43 years, mean 45 years, and range 0-100 years. Thus, 7.5% (1,935/25,948) of males, and 4.4% (1,980/44,569) of females tested were positive. There was a statistically significant difference in these proportions (z = 16.8595, p<0.01, calculated with Z-test). The difference appeared to decrease towards the end of the study period (Figure 4).

**Figure 4.**
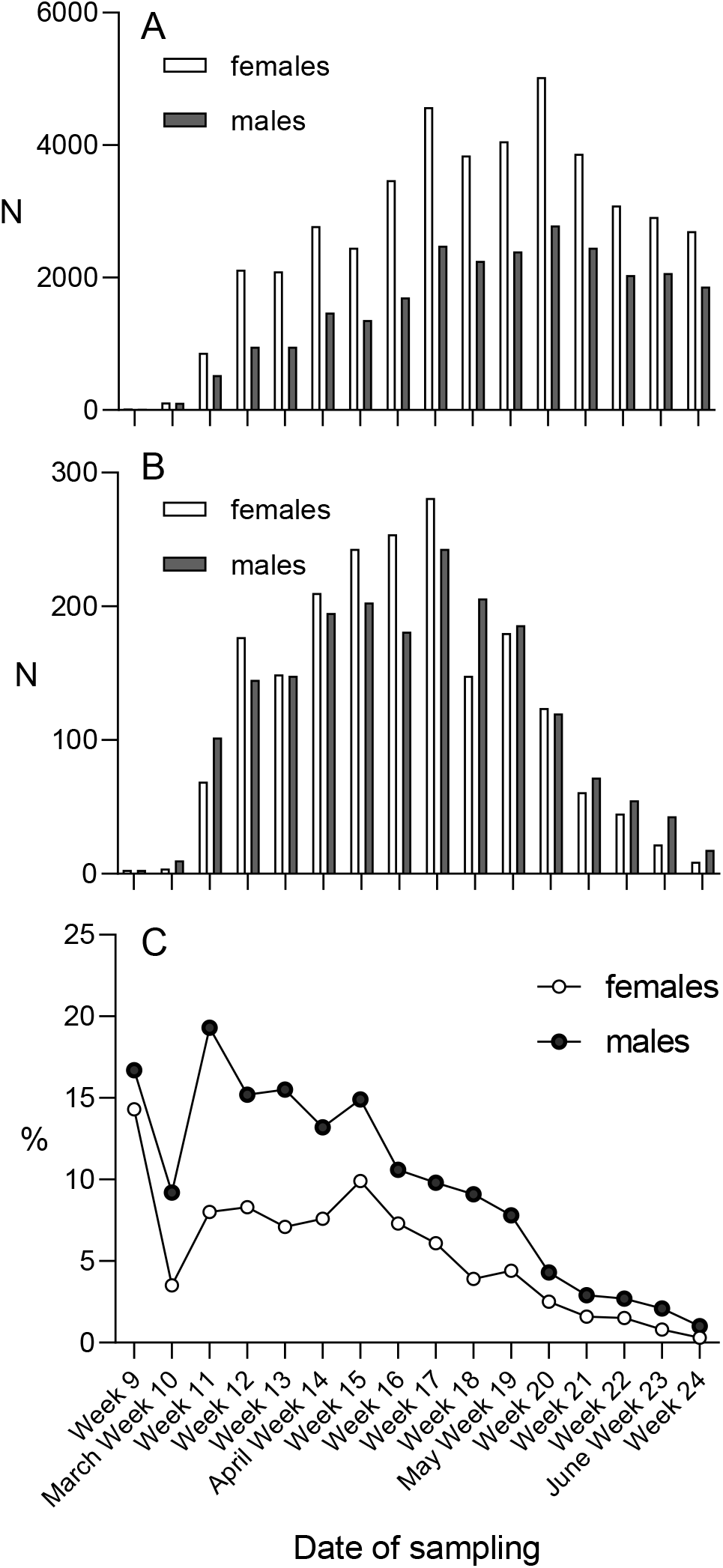
A) The sex-specific number of SARS-CoV-2 individuals tested for SARS-CoV RT-PCR by calendar week of 2020. B) The sex-specific number of SARS-CoV-2 positive cases by calendar week of 2020. C) The sex-specific proportion (%) of SARS-CoV-2 positive cases by calendar week of 2020.

## Discussion

By 30 June 2020, 80.2% of all tests performed, and 77.3% of new positive cases detected within the Hospital District of Helsinki and Uusimaa were analysed in HUSLAB [7], rendering our surveillance data representative of Greater Helsinki. In Finland, COVID-19 has mostly affected the Greater Helsinki area, which represents 30% of the Finnish population [2]. Indeed, 73% of the positive cases in Finland by the end of June 2020 have been detected in Greater Helsinki [7].

While the number of test positive cases was similar in males and females, the positivity rate in males was significantly higher than in females in our study. This may, to a small degree, be explained by the overrepresentation of healthcare workers and female predominance in the elderly. However, our data suggest that men pursued SARS-CoV-2 testing much less frequently than women. Consequently, a subset of COVID-19 infections in men may have gone undetected.Abundant evidence shows that women generally seek more health care services than men [8,9].

The proportion of young adults in positive cases increased towards the end of the study period, which may suggest their returning back to social behaviour with risk of infection. Extension of testing criteria into mild symptoms may have also contributed. Some of the restrictions imposed due to COVID-19 in Finland were lowered on 1 June, including opening of restaurants [10]. However, considering the incubation period [11], it does not appear likely that relaxing restrictions played a role with this observation.

The high incidence rate observed in the elderly is in line with earlier reports [12]. Vigilance in the infection control in long-term care facilities remains cricitally important in the mitigaton of the pandemic.

As to limitations, our data did not include all tests analysed in Greater Helsinki. No clinical data were available. A bias may have been introduced by the restricted sampling criteria during March 2020. During the first weeks, control swabs were recommended for the positive cases – a policy since abandoned. Also, during the first two weeks of April less preferred oropharyngeal swabs were used due to the global shortage of nasopharyngeal swabs, which may have temporarily influenced the overall test sensitivity.

Large dips in testing frequency were observed on every weekend, and also during public holidays (Figure 1). Simultaneously, the number of new positive cases dropped each time, and the epicurve (Figure 1b) may suggest that this testing deficit was not fully compensated during the following weekdays.

In a response to need for large-scale testing, HUSLAB switched from two-shift work into a three-shift work on 23 March, and personnel was reallocated. A laboratory-developed test [3] was ready in use in mid-January, and testing on the cobas® 6800 system late March. Through constant global shortages on reagents and plasticware, the need to deploy several independent methods was evident to secure laboratory services and capacity building. The Amplidiag® COVID-19 tests were deployed as of mid-April, and the Novodiag® sample-to-answer test as of mid-May.

In preparation for the potential next epidemic waves of COVID-19 pandemic, the difference in health-care seeking behaviours between men and women needs to be accounted for to facilitate high universal testing frequency. This is particularly relevant as men are at higher risk of fatal infection [13], more likely to be hospitalised [14] and admitted to ICU [15] due to COVID-19. In addition, advocating and maintaining social behaviours that reduce risk of infection remains a key measure of mitigation in all age groups.

## Data Availability

Data is available upon request.

## Acknowledgements

We would like to thank the staff at the Department of Virology and Immunology working with SARS-CoV-2 diagnostics.

## References

1. World Health Organization (14 April 2020). Strategic preparedness and response plan. Retrieved from https://www.who.int/publications/i/item/strategic-preparedness-and-response-plan-for-the-new-coronavirus

2. Official Statistics of Finland (OSF): Population structure [e-publication]. ISSN=1797-5395. Helsinki: Statistics Finland [referred: 28.6.2020]. Access method: http://www.stat.fi/til/vaerak/index_en.html. Accessed 27 mJune 2020.

3. Corman VM, Landt O, Kaiser M, Molenkamp R, Meijer A, Chu DKW, et al. Detection of 2019 novel coronavirus (2019-nCoV) by real-time RT-PCR. Euro Surveill 2020; 25(3).

4. Mannonen L, Kallio-Kokko H, Loginov R, Jääskeläinen A, Jokela P, Antikainen J, et al. Comparison of two commercial platforms and a laboratory developed test for detection of SARS-CoV RNA. Manuscript submitted.

5. Jokela P, Jääskeläinen AE, Jarva H, Holma T, Ahava MJ, Mannonen L, et al. SARS-CoV-2 sample-to-answer nucleic acid testing in a tertiary care emergency department: evaluation and utility. Manuscript submitted.

6. Haveri A, Smura T, Kuivanen S, Österlund P, Hepojoki J, Ikonen N et al. Serological and molecular findings during SARS-CoV-2 infection: the first case study in Finland, January to February 2020. Euro Surveill 2020;25(11):2000266.

7. National Institute for Health and Welfare, THL. National Infectious Disease Register. Retrieved from: https://sampo.thl.fi/pivot/prod/fi/epirapo/covid19case/fact_epirapo_covid19case

8. Deveugele M, Derese A, van den Brink-Muinen A, Bensing J, De Maeseneer J. Consultation length in general practice: cross sectional study in six European countries. BMJ. 2002;325(7362):472.

9. Thompson AE, Anisimowicz Y, Miedema B, Hogg W, Wodchis WP, Aubrey-Bassler K. The influence of gender and other patient characteristics on health care-seeking behaviour: a QUALICOPC study. BMC Fam Pract. 2016;17:38.

10. Prime Minister’s Office in Finland. Government decides on plan for hybrid strategy to manage coronavirus crisis and for gradual lifting of restrictions. Retrieved from https://vnk.fi/-/hallitus-linjasi-suunnitelmasta-koronakriisin-hallinnan-hybridistrategiaksi-ja-rajoitusten-vaiheittaisesta-purkamisesta?languageId=en_US

11. Lauer SA, Grantz KH, Bi Q, Jones FK, Zheng Q, Meredith HR, Azman AS, et al. The Incubation Period of Coronavirus Disease 2019 (COVID-19) From Publicly Reported Confirmed Cases: Estimation and Application. Ann Intern Med 2020;172:577–582.

12. Natale F, Ghio D, Tarchi D, Goujon A, Conte A. COVID-10 Cases and Case Fatality Rate by age. Retrieved from: https://ec.europa.eu/knowledge4policy/publication/covid-19-cases-case-fatality-rate-age_en

13. The Novel Coronavirus Pneumonia Emergency Response Epidemiology Team. The Epidemiological Characteristics of an Outbreak of 2019 Novel Coronavirus Diseases (COVID-19) — China, 2020. China CDC Weekly, 2020, 2(8): 113–122.

14. Garg S, Kim L, Whitaker M, et al. Hospitalization Rates and Characteristics of Patients Hospitalized with Laboratory-Confirmed Coronavirus Disease 2019 — COVID-NET, 14 States, March 1–30, 2020. MMWR Morb Mortal Wkly Rep 2020;69:458–464.

15. Grasselli G, Zangrillo A, Zanella A, et al. Baseline Characteristics and Outcomes of 1591 Patients Infected With SARS-CoV-2 Admitted to ICUs of the Lombardy Region, Italy. JAMA. 2020;323(16):1574–1581.

